# Frontline healthcare workers’ mental health and wellbeing during the first year of the COVID-19 pandemic: Analysis of interviews and social media data

**DOI:** 10.1101/2022.04.29.22274481

**Authors:** Norha Vera San Juan, Sam Martin, Anna Badley, Laura Maio, Petra C. Gronholm, Caroline Buck, Elaine C. Flores, Samantha Vanderslott, Aron Syversen, Sophie Mulcahy Symmons, Inayah Uddin, Amelia Karia, Syka Iqbal, Cecilia Vindrola-Padros

**Affiliations:** Rapid Research Evaluation and Appraisal Lab (RREAL), University College London, UK; Institute of Psychiatry, Psychology and Neuroscience. King’s College London. UK; London School of Hygiene and Tropical Medicine; University of Oxford; Trinity College Dublin

**Keywords:** Mental health, frontline, healthcare workers, COVID-19, health services research, LISTEN method

## Abstract

**Background:** The COVID-19 pandemic has shed light on the fractures of healthcare systems around the world, particularly in relation to the healthcare workforce. Frontline staff have been exposed to unprecedented strain and delivering care during the pandemic has impacted their safety, mental health and wellbeing. Rapid Research methods and big qualitative data offered a unique opportunity to gain insight into perceptions and experiences during this time.

**Objective:** The aim of this paper was to explore the experiences of Health Care Workers (HCWs) delivering care in the UK during the COVID-19 pandemic to understand their wellbeing needs, experiences and strategies used to maintain wellbeing at individual and organizational levels.

**Methods:** We analysed 94 telephone interviews with HCWs and 2000 tweets about HCWs mental health during the first year of the COVID-19 pandemic applying Collaborative and Digital Analysis of Big Qualitative Data in Time Sensitive Contexts (LISTEN).

**Results:** Results fell under six themes: redeployment, clinical work, and sense of duty; wellbeing support and HCW’s coping strategies; negative mental health effects; organisational support; social network and support; and public and government support. Redeployment generated anxiety mainly due to limited prior training and risk assessments, and the barriers of adapting to a new working environment while wearing PPE. HCWs struggled to access wellbeing support due to time constraints. In terms of ill mental health, mentions of feelings of trauma, PTSD and anxiety were prominent. HCWs’ mental health was particularly affected by the copious amount of bad news on media and at home and the fear of infecting their loved ones.

**Conclusions:** These findings demonstrate a need for open conversations, where staff wellbeing needs and the strategies they adopted can be shared and encouraged, rather than implementing solely top-down psychological interventions. At the macro level, findings also highlighted the impact on HCW’s wellbeing of public and government support, as well as the need for ensuring protection through PPE, testing, and/or vaccines for frontline workers.

## INTRODUCTION

High levels of stress, burnout, and symptoms of poor mental health have been well known among healthcare workers (HCWs) for a number of years [1]. Though many health systems include mechanisms to support HCW’s wellbeing, the COVID-19 pandemic exacerbated the fractures of systems around the world in relation to protecting their healthcare workforce. Frontline staff, in particular, have been exposed to unprecedented strain. Delivering care during the pandemic impacted their own, and their loved one’s safety, and complexities of the context placed them in hard and uncomfortable situations for which they had not necessarily been trained [2-4]. Studies showed the toil on physical and mental health of delivering care over long hours, under the heat of full personal protective equipment (PPE), the pressures of changing guidelines and the rising rates of infection among HCWs [5-8]. This anxiety was exacerbated by the high rates of hospital deaths, and the added responsibility many HCWs felt to accompany patients during the last moments of their life so they would not die alone [9]. Dowrick et al [10] pointed to the significant emotional labour involved in affective practices to mitigate the limitations arising from physical distancing, such as maintaining communication with patient families, and keeping in touch with work colleagues who were also going through a difficult time. While infection control measures were crucial for limiting the spread of COVID-19, they required complex additions to HCWs workload in order to reorganise necessary interactions at work. Our previous work on this topic additionally highlighted distancing measures completely disrupted environmental factors that influence HCWs wellbeing [4], such as leisure time. Not only does the emotion involved in delivering care under these circumstances add copious amounts of strain on HCWs, but the moral injury of caring for patients under time and resource constraints, has also been a frequently mentioned factor associated with poor mental health, since the start of the pandemic [11].

Nursing staff, in particular, have been severely impacted by the pandemic [12, 13]. Some authors have argued that nursing staff have experienced moral conflicts and complex ethical issues due to the need to make difficult decisions in the context of medical rationing produced by a high patient demand and limited resources [14, 15]. Nurses have expressed fears of becoming infected and transmitting the virus to family members [16]. Several studies have reported higher rates of work-related stress among nurses, a higher rate of burnout among nurses when compared to medical staff, while being female and a nurse have both been identified as risk factors for poorer mental health outcomes [17, 18]. Other risk factors for psychological distress reported in the literature include being younger, more junior, being the parent of dependent children, having infected family members or having a frontline role [19-21].

Evidence from previous emergencies, including previous epidemics, has pointed to the detrimental effects of working in these conditions and has highlighted the importance of considering frontline workers’ mental health and well-being [22, 23]. During the COVID-19 pandemic, efforts were made to integrate this evidence and learning from previous epidemics in the development of wellbeing guidelines and support interventions, but many of these guidelines and interventions focused mainly on the assessment of clinical outcomes, such as the identification of symptoms of PTSD [24, 25]. Emerging evidence is starting to show that social and organisational measures such as maintaining clear communication, providing adequate personal protective equipment (PPE) to staff, allowing staff to have adequate breaks and rest and delivering psychological support in a timely and practical way could help reduce the risk of adverse mental health outcomes and improve the wellbeing of staff [20, 21, 24, 26].

In an early analysis of the mental health needs of HCWs during the COVID-19 pandemic in the UK - we found that support guidelines did not implement a holistic depiction of wellbeing. Even though some guidelines have recently been expanded to consider the contextual factors that might be shaping wellbeing, there is limited evidence on our understanding of the wellbeing of HCWs (beyond individual mental health) in the context of complex health emergencies such as the COVID-19 pandemic.

The aim of this paper was to conduct an in-depth exploration of the experiences, needs, and wellbeing strategies of HCWs and Institutions delivering care in the UK during the COVID-19 pandemic. We did not limit our analysis to clinical mental health and defined wellbeing following the SPICE model [25] that considers social, political, and quality of life factors. We also explored how these experiences changed during different stages of the pandemic.

## METHODS

This study is part of a larger ongoing project led by the Rapid Research, Evaluation and Appraisal Lab (RREAL) which was designed as a qualitative rapid appraisal with the aim of analysing healthcare workers’ experiences and perceptions of delivering care during the COVID-19 pandemic [7]. Rapid appraisals are developed to collect and analyse data in a targeted and iterative way within limited timeframes, and to ‘diagnose’ a situation. The study combined telephone interviews with a purposive sample of HCWs based in the UK and the analysis of social media data.

### Data collection

#### Interviews with HCWs

The authors assert that all procedures contributing to this work comply with the ethical standards of the relevant national and institutional committees on human experimentation and with the Helsinki Declaration of 1975, as revised in 2008. This study was approved by the Health Research Authority (HRA) in the UK (IRAS: 282069) and the local Research and Development Offices (R&D) where the study took place. All participants provided consent before taking part.

Semi-structured interviews were conducted over the phone by members of the RREAL research team, including (AD, LM, KH, SMU, CVP, KS, GS, AS, LJM, HF, NR, CB, SLJ). We report on the findings from 94 interviews carried out between March 2020 and March 2021. The interview topic guide was revised four times during this period, following emerging findings and changing public and policy concerns at the different stages of the pandemic. A detailed description of data collection and sampling methods can be found in Vera San Juan et al [25].

A group of the authors (NVSJ, LM, CB, PCG, AB, AS, EF, IU, SMS) prepared interview data for analysis by performing selective transcription of extracts from the interviews and interview notes that were related to mental health and wellbeing as previously defined [4]. This team includes health service researchers, academics, nurse practitioners, anthropologists, and physicians.

#### Social media data

Data were collected from social media accounts of self-identified HCWs in the UK (Appendix 1) in the periods between March 19^th^ 2020–March 19^th^ 2021. Data analysis was guided by the collaborative coding framework analysis provided by the interview analysis. The key themes produced from the interview framework were used to inform the metadata and key search terms of social media data collection. As a result of this, advanced Boolean search terms were created that reflected keywords from the themes in the collaborative coding framework (Appendix 1). These Boolean search terms were then used to mine Twitter data archives using the media monitoring Meltwater software (TM, 2020) for English language tweets related to the themes found in interview data (and that were engaged with (liked/quoted/retweeted) more than once). An initial dataset of 775k tweets were sampled from posts shared by HCWs in the UK and were then filtered and cleaned using the inclusion and exclusion criteria, during the later social media analysis stage.

### Data synthesis and analysis

#### Interviews with HCWs

Interviews were analysed by conducting a Collaborative and Digital Analysis of Big Qualitative Data in Time Sensitive Contexts (LISTEN). This analysis consists of iterative cycles intercalating team discussion and the use of digital text and discourse analytics tools to analyse related social media data (see Figure 1).

**Figure 1.**
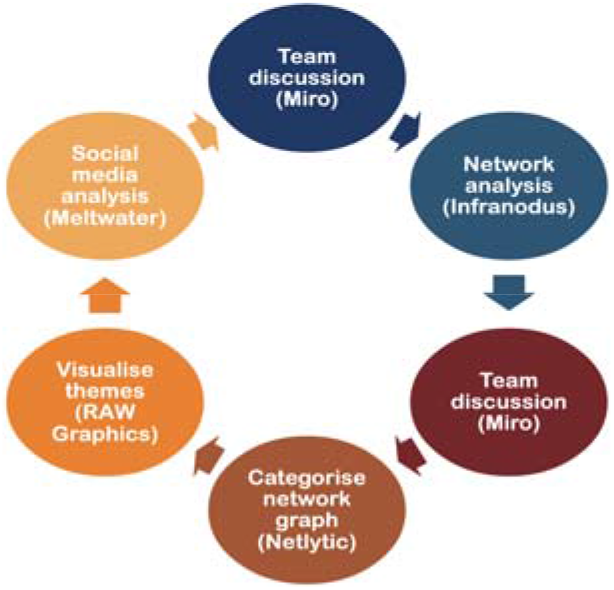
LISTEN analysis cycles

First, the group of authors who prepared the data for analysis held a guided group discussion (see Collaborative Matrix Analysis [27]) using the Whiteboard Tool Miro (Desktop, version 0.42) to outline: (1) the main emerging themes; (2) patterns in the presence of themes in interviews with specific population or professional groups; (3) negative cases, these are cases that appeared to be out of the norm. Based on this, a preliminary coding framework was developed and shared with the digital analysis team (SM and SV) who, using the Infranodus discourse analysis tool [28] developed an analytical coding framework which predicated a preliminary scan of the data, group discussions and thematic discourse mapping. They conducted a digital analysis of co-occurring themes with betweenness centrality and frequency analysis [29, 30], and inputted it in a Microsoft Excel matrix. The framework was refined during team discussions and doing further digital inquiries using big qualitative keyword analytics to test sample data with Netlytic [31].

Results from the digital analysis were shared with the research team and a joint team discussion was held to refine and finalise the coding framework. Codes were defined in a coding book and inputted into a coding matrix on Excel that included codes and participant characteristics in the columns, and cases in rows. Researchers completed the matrix for sets of interviews that they were assigned. NVSJ cross-checked the data during the coding process to ensure consistency across researchers.

Once data indexing was complete, codes were divided among the team to be synthesized, and selected quotes from the interview transcripts were chosen to exemplify themes. We then held a third team discussion using Miro to assess links between key topics emerging within each code, based on this, we developed the final set of themes encompassing the main issues raised by frontline staff. NVSJ reviewed the definitions of the themes, alongside crosschecking with the thematic discourse mapping conducted on Infranodus [28], to ensure consistency in relation to the grouping of codes and the selection of illustrative quotes.

#### Social media data

First, tweets were screened to assess data quality and alignment with inclusion criteria. The process to complete this consisted of 5 steps: (1) The resulting social media data (tweets) that were collected were then analysed and coded using the key themes and keywords from the interview-based collaborative framework as a guide. (2) SM and SV selected anonymized tweet examples [32] that could exemplify mental health themes and patterns of discourse in the data. (3) The results from this analysis were then used to structure a comparative social media analysis and sentiment analysis framework (Appendix 3). (4) An Include/Exclude criterion was developed within our social media analysis framework to maintain good data quality, paying attention to the importance of using a mixed methods approach, which involves incorporating both qualitative and quantitative evaluations to assess data quality [33].

Nine independent coders annotated the tweets (NV, SM, SV, CB, ECF, AS, IU, PCG, LM), allocating them into the five key themes from the social media analysis coding framework (Appendix 3). Annotation was completed in 2 rounds, where the coders met to discuss and agree on intercoder-reliability. After meeting and discussing the tweets that they had queries about, the annotators were able to agree on the coding of all remaining posts.

#### Sentiment analysis

We aimed to get a better understanding of sentiment of tweets within the context of the HCW experience by creating a manual sentiment framework. By drawing on previous research [30, 32], we set up our own framework with definitions based on our analysis, using the more specific qualitative lens, and carefully constructed the qualitative framework. This drew upon part 4 of the LISTEN cycle, where we created a sentiment framework that was informed by an analysis of the themes from the qualitative interview coding framework in this study. Sentiment was measured in terms of differences in attitudes towards the impact of mental health on the individual, as well as impact on delivery of care within the context of the pandemic (Appendix 2). Posts or articles were classified as positive towards individual mental health experiences of work, such as redeployment, wellbeing, organisational, social/family, public or government support on the frontline if, for example, they were affirming of mental health support/experience or communicated overall trust of hospital or government guidelines. Posts were marked as negative if they contained negative attitudes or arguments against the way the mental health of HCWs was supported, shared bad experiences, or discouraged the following of government or hospital guidelines. Posts were then marked as neutral if they contained only a general statement, with no expression of sentiment or opinion. The text of tweets was analysed in terms of discourse content, sentiment analysis and frequency per topic.

#### Discourse analysis

Thematic discourse mapping was used to analyse the social media data within each of the five coding categories (Appendix 3), using the software Infranodus [28]. This analysis was used to map co-occurring themes, keyword frequencies and patterns occurring in the data with regard to discussions about mental health experiences of HCWs, as well as the strength of the betweenness centrality of sub-themes (connections between sub-themes that link different types of conversation clusters together) [34].

## RESULTS

### Interview characteristics

We extracted information relevant to wellbeing and mental health from 94 interviews with professionals across London including nurses, physiotherapists, anesthetists, and consultants. We analysed interviews between March and September 2020, that lasted between 12 minutes and 2 hours (37 minutes on average). Professionals varied in years of experience and grades ranging from recently qualified (less than 1 year) to professionals with over 30 years of experience. Around 66% of the sample was female and the average age was 38 (22-60 range). A more detailed description of the sample and interview characteristics can be found below (Table 1-4).

**Table 1.** Gender demographics of participants

| Gender | No. | in % |
| --- | --- | --- |
| **Female** | 62 | 66% |
| **Male** | 27 | 29% |
| **Not known** | 5 | 5% |
| **Grand Total** | 94 | 100% |

**Table 2.** Age demographics of participants

| Age | Column1 |
| --- | --- |
| **Min** | 22 |
| **Max** | 60 |
| **Average** | 38.2 |

**Table 3.**
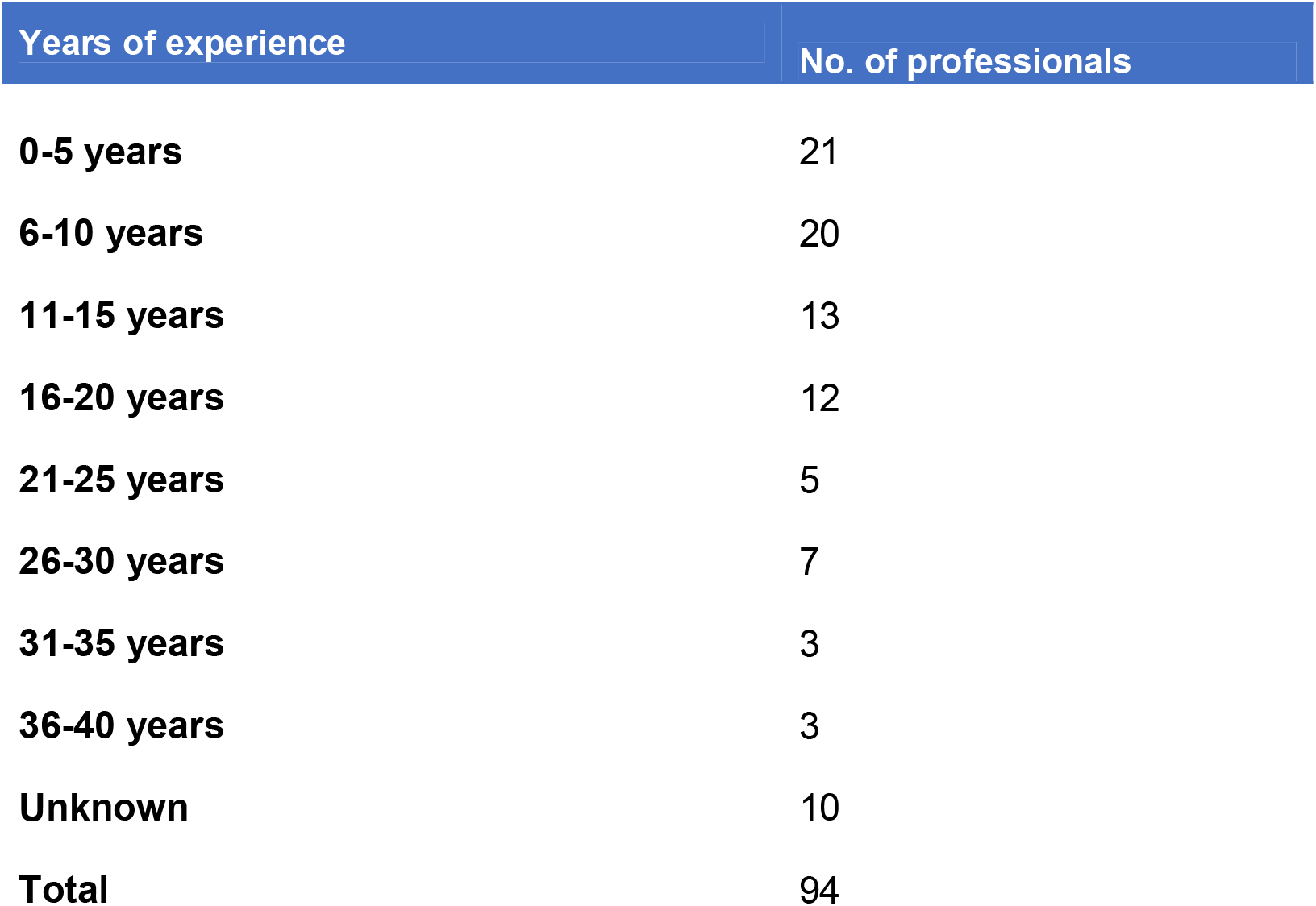
Years of experience

| Years of experience | No. of professionals |
| --- | --- |
| **0-5 years** | 21 |
| **6-10 years** | 20 |
| **11-15 years** | 13 |
| **16-20 years** | 12 |
| **21-25 years** | 5 |
| **26-30 years** | 7 |
| **31-35 years** | 3 |
| **36-40 years** | 3 |
| **Unknown** | 10 |
| **Total** | 94 |

**Table 4.** Ethnicity demographics of participants

| Ethnicity (macro categories) | No. |
| --- | --- |
| **Asian/British Asian** | 7 |
| **Black/Black British** | 3 |
| **Mixed** | 2 |
| **White/White British** | 36 |
| **Not known** | 46 |
| **Grand Total** | 94 |

### Social media data characteristics

From a total of 775k tweets, quoted tweets and replies - there were 104k original tweets, which were quoted/discussed 192k times, replied to 36.4k times, and retweeted 443k times. 19k commented on and quoted tweets/retweets and 36.4k were replies to original tweets/retweets. Screening for the inclusion/exclusion criteria (Appendix 4) resulted in 2000 tweets, which were included in the final analysis. Comparing the final agreed coding after discussions with the annotators’ initial coding, the accuracy of the individual annotators was 71% for the themes (F-score 0.62), and 81.4% for the Includes and Excludes (F-score 0.76).

### Main themes

The results present experiences that were perceived by HCWs as affecting their wellbeing; expressions of wellbeing needs; and wellbeing strategies presented by staff. Findings from the analysis of interviews and social media data fell under six themes: redeployment, clinical work, and sense of duty; wellbeing support and HCW’s coping strategies; negative mental health effects; organisational support; social network and support; and public and government support. We describe each theme and present illustrative quotes in the following subheadings.

#### Redeployment, clinical work, and sense of duty

Whilst staff often willingly volunteered to be redeployed, the prospect caused anxiety for a number of HCWs due to speculation that new roles might be entirely outside of normal scope of practice. Some staff reported that options for redeployment were offered via online surveys, giving little description of what they were signing up to do. The pace of the crisis required rapid decision-making amid vague details of what the new role might entail. For team managers, being unable to provide clarity around who would be redeployed and when was also cause for distress.

> “*We saw other people were very anxious beforehand and we did a lot of work around trying to support people around those anxieties”* (C0V40, Consultant Pediatrician).

New location, unknown faces, difficulty communicating while wearing PPE, and rapidly learning different ways of working were reported as challenging aspects about navigating redeployment. Concerns about being exposed to the virus were high and participants informed there was no risk assessment of vulnerable HCWs or those with susceptible family members were undertaken, particularly early on in the pandemic. Additionally, typical mechanisms that support a change of job (e.g., induction/orientation, management guidance) were limited, increasing the difficulty of day-to-day working. This was particularly true for those redeployed to a role with little relation to their normal job remit, (e.g., allied health professionals reassigned from outpatient/community settings to working with inpatients on hospital wards).

> “*In terms of just levels of stress, I think for me it’s been very difficult (to) distinguish between not just purely COVID but then also being in a new job, a different role… I know for sure I wouldn’t have felt this stressed had I been in somewhere new (where) I knew the people…I think that’s added a massive amount to me*” (COV52, Charge Nurse).

Redeployment was not always voluntary. Some staff reported that they were selected without having much say in the matter, with department managers forced to make the selection. Other participants recounted how an individual’s suitability to a particular discipline was not taken into consideration, especially when reassigning a member of staff to a complex environment such as the ICU.

> *“I think you have some members of staff that were redeployed (who) felt that they were in some way singled out. Why did they have to go? Because it wasn’t necessarily people that volunteered”* (COV100, Nurse Manager).

Whilst a number of participants expressed relief and some pride that they were not redeployed (their roles were deemed important for the maintenance of certain services (e.g., surgery, cancer treatment), some participants expressed feelings of shame at avoiding redeployment (intentionally or otherwise) amid the increasing narrative of ‘heroism’ circulated by the media.

> “*Guilty that we were not doing more … that’s why a few of my colleagues volunteered … to do other things in a training capacity or clinical service … that is how we assuaged our guilt*” (COV57, anesthetist).

Some HCWs who had experienced a positive redeployment shared their pride at being able to help, but also looked ahead to return to their previous roles:

> *“Today, my redeployment to Covid Intensive Care came to an end. I have never been so proud to have been part of such an incredible workforce that have given and continue to give their absolute all to deliver the best care possible. Next stop > 11th floor in Stroke* □*”* (COVTweet423, Physiotherapist)

Feelings of resentment were reported as surfacing once some services returned to normal, with the inequity between those redeployed and those who stayed in their normal role becoming apparent, especially if redeployed staff had not been particularly busy, as was the case for sexual health services

> “*There were lots of people I think that have now come back that are finding it difficult to fit back into the team. There’s quite a lot of resentment there*” (COV100, Nurse Manager, Sexual Health).

Nurses were perceived by other professionals as enduring more challenges than other disciplines. While team leads in general expressed keeping track of team wellbeing was an important challenge, nurse leads in particular reported struggling to train new staff and manage their physical and emotional wellbeing while keeping on top of their increased clinical workload.

> *“You have these really frightened inexperienced nurses who sometimes haven’t seen a patient die ever or for a long time who are terrified and immobilised” (*COV73, Lead Nurse).

Changes to patient demand and complexity led to many HCW’s sharing their discomfort and distress at not being able to provide the level of care and patient experience that they would like and “forced to be comfortable with the uncomfortable” (COV95, Profession). Allied Health Professionals described how they were required to juggle clinical work from their normal role whilst simultaneously being redeployed to a new position, resulting in neither role being performed optimally:

> *“There was a lot of pressure on us to help everybody … we were finding it difficult ourselves”* (COV90, ICU dietician).

These feelings of exhaustion were enhanced by levels of poor patient outcomes and not being able to socialise with colleagues. HCW’s described a constant battle between the relentless demand for clinical care and needing to rest.

> *“So much is asked of staff who are already stretched and underpaid. Worn out and stressed staff can’t cope”* (COV96, Critical care dietician).

Overall, a feeling of sadness underpinned many HCW’s experiences. This was often related to caring for people who were dying in the absence of family members. HCW’s spoke of absorbing the sadness of dying and the amount of death as well as trying to provide care and consolation to absent relatives remotely.

> *“I think the hardest thing is, is that, (…) they cannot die alone. That’s so incredibly hard thing to come to terms with, isn’t it? You know, it’s… but, you know, we never allow anyone to die alone, we were always in there with them, because that’s what you do as a nurse”* (COV46, Infection control nurse).

### Wellbeing support and HCW’s coping strategies

Most staff believed that the pandemic led to strong leadership from management and facilitated more compassion and awareness to mental health and well-being. Twitter was used as a place to share practical guidelines and wellbeing resources.

> *“A guide to support managers to strike the right balance between directive and compassionate leadership that will help to ensure their teams come through the #COVID19 crisis with a greater resilience and mutual respect. Both critical to the recovery phase”* (COVTweet1269, Chief Nursing Officer).

HCWs were offered a range of mental health and wellbeing support including appointments with clinical psychologists, emails with wellbeing recommendations, and peer support group meetings. HCWs who interacted with psychologists found it useful and expressed their gratitude for the opportunity to be in contact with such professionals. Staff felt it was a privilege to have access to a wide range of support, however, many expressed it had not been possible to make use of these services. Lack of time due to staff shortages, no holidays, and sense of duty were often highlighted as barriers for HCWs to access support.

> *“I think the problem is being able to access them on a shift, when it’s just so busy and manic. It’s one thing having the space to go and relax, but if you haven’t got the time to go and do it, then, that’s a different thing”* (COV52, ICU Nurse).

Some HCWs described their experience as “groundhog day-like”, having no sense of progress or direction. Managers and more experienced ICU clinicians were central to providing guidance and support despite having significant additional clinical work burdens.

> *“Definitely, definitely on sleeping, definitely on mental health because it was constant stress, constant worry, you know as I say as a general manager people would come to me for assurances and I wasn’t able to give the assurances that people were looking for and it was up to me to try and stay calm and support my staff but the same time I felt very little support in return from my superiors and the rest of the organization”* (COV117, General Manager, Critical Care).

Some managers put in place practices to mitigate this, such as enforcing breaks, or being available to talk about things beyond work. This provided some space for staff to briefly focus on their well-being. However, there were dilemmas about whether these wellbeing practices should be mandatory.

> *“I can’t make it mandatory, but I strongly encourage people to have clinical supervision*… *You’ve got that offer there, you’re an adult, you’re a professional, you have to take some responsibility for your own health and wellbeing”* (COV73, Lead nurse).

Regarding support from peers, infection control rules meant staff could not socialise in ways they were used to both inside and outside of hospitals. HCWs missed informal interactions, particularly staff working remotely and teams who were broken up due to redeployment. However, most staff working in new environments found it invigorating. Team morale was reported as high, with some people thriving on change, learning new skills, and strengthened relationships and respect amid the difficult times shared. Reduced hierarchy also played into the sense of togetherness and team spirit.

> *“I think there’s been a lot of like solidarity between the teams, trying to get through this and coming together and appreciating the help we’ve had from other disciplines, the redeployed staff, so hopefully they’ve seen how hard we work, and maybe appreciate the efforts of ICU more. I think there’s definitely been a good coming together and an ethos of like we’ve got to try and do our best to get through this”* (COV52, ICU Charge Nurse).

Furthermore, some staff felt they did not need to access support due to their existing knowledge about mental health and techniques they knew to build resilience. Support from family and friends also played a key role in keeping a positive mindset. Staff incorporated their own coping strategies in maintaining their wellbeing such as using mindfulness apps, performing exercise and being outdoors, which allowed them to disconnect during times when they were feeling overwhelmed and exhausted.

> *“Personally feel like I already had enough teaching, and I was prepared for that, it wasn’t hugely beneficial for me, but for other people it was”* (COV43, GP trainee).

### Negative mental health effects

Participants described a range of negative mental health effects. Many reported feelings of trauma and PTSD-like symptoms, along with a sense of being overwhelmed, emotional, and physical exhaustion. In addition to distressing factors mentioned earlier, HCWs were affected for example by the onslaught of news, and negative events occurring to relatives and close friends.

> *“What I was witnessing over the past, in April anyway, was some of the most distressing stuff I’ve ever seen professionally”* (COV65, Palliative Care Consultant).

HCWs also discussed their experience of caring for coronavirus patients in different “Covid Zones” within Covid Wards (red, orange, green and blue), while some expressed anxiety about ICU capacity, the prolonged amount of time nurses spent working in these wards, and the extreme risks taken with varying access to adequate PPE. Others also reported being in charge of Covid ICUs, where staff were busy, overwhelmed, and depleted by illness, exhaustion, and also the need to quarantine. Some HCWs expressed during the height of the pandemic they actively did not stop to consider potential negative effects on their mental wellbeing. Often, HCWs spoke of potential trauma that was not fully registered until later, at which point many struggled to fully process their experiences and reported feeling less resilient than before.

> *“I think now we’ve got a bit more space to sort of process because I think when it was all happening it was* … *you just went into almost auto pilot and you couldn’t really emotionally process anything anyway”* (COV108, Speech and language therapist).
>
> *“I don’t know whether it’s sort of had an impact on my attention or anything but it’s definitely, erm, I’m feeling more* … *I don’t feel as robust as I did”* (COV108, Speech and language therapist).

A key source of anxiety was the threat of becoming infected with Covid-19. This fear was exacerbated by coming into contact with Covid-19 patients, seeing severely unwell and dying patients, and, at the beginning of the pandemic, the lack of understanding about the illness and limited insights regarding possible treatments. Some staff members also spoke of how their anxiety increased as time went on and the full extent and severity of the pandemic became apparent.

> “*Obviously it makes you fear for yourself and family and friends and those sorts of things and especially with headlines that lots of healthcare workers are you know getting sick and then their dying obviously kinda adds to those concerns as well*” (COV69, Senior PT).

Anxiety was also reported in relation to fears of inadvertently infecting others. This was a worry particularly for those who lived with or cared for vulnerable family members, such as elderly parents. Other sources of anxiety related to Covid-19 infections were concerns for colleagues who became unwell, awareness of the burden placed on family members who worried about frontline staff, worries for the general public who lacked medical knowledge in dealing with the pandemic. For some, anxiety was brought on by other work-related factors, such as having to give up usual clinical work, lack of control, and not being able to switch off from work. An example of such anxiety was expressed in a tweet by a doctor who had worked on the frontline during the first wave of the pandemic.

> *“*…*extremely vulnerable and sad, and did have a brief mental health relapse. I had to seek help through my GP and saw a psychologist. Why did I become unwell? Stress surrounding lack of PPE, re-deployment, worries about catching COVID-19 and bringing the virus home to my family*…*”* (COVTwitter845, Doctor).

An adverse mental health impact was issues with sleep. Racing thoughts, constant exposure to negative events and worries and increases in number of night shifts, led to disrupted sleep, which contributed to further exhaustion.

> *“It was a recurring theme where I’d say I’d been awake since four, I couldn’t get back to sleep after I woke up feeling this overwhelming feeling of threat, panic, but not, not like palpitations or breathlessness just an impending doom, that was kind of what it was*…” (COV95, Consultant Anesthetist).

Another key mental health impact was the feeling of sadness over witnessing a high number of deaths. Participants also spoke of how it was particularly difficult to see the impact of Covid-19 on patients who would normally not be expected to become severely unwell, such as otherwise healthy younger people.

> *“Some people that stick in my head are patients that were quite young erm, like patients in their thirties or patients in their forties with zero comorbidities coming into it, and then were in the hospital for about fifty days and had trouble weaning from the ventilator*” (COV83, PT).

Many also described worries about what was ahead. A key source of concern was fear of further Covid-19 waves, after first-hand experience of how challenging the first wave was and how frontline workers were still reeling in its aftermath. Many spoke of concerns regarding the suspended non-emergency procedures and treatments which still needed to be performed alongside routine care by a jaded workforce. Participants also sensed that many patients had not approached health services with their concerns due to ‘stay at home’ guidance and/or a preference to avoid hospitals, which were seen as Covid-19 hotspots. This meant health problems had likely worsened over time and would now require more attention.

> *“But obviously there is a huge amount of people that just aren’t going to be seen that are just still waiting on a waiting list and our expectation is that you know there’s going to be new patients that are still gonna probably need to be referred so that when we are allowed to open up we are going to have a huge backlog to go through plus potentially more patients that are going to need to be seen afterwards and that is going to be a challenge within itself* “ (COV69, Senior PT).

These concerns and causes for distress were also present in the overall sentiment (Appendix 2) of tweets shared about HCWs individual mental health. One of the key topics of concern was the exhaustion felt by both doctors and nurses, with some reporting feeling utterly shattered after several tough weeks working on the frontline looking after Covid patients. Nurses shared reports of being signed off work with anxiety, while some doctors saw several family members admitted into Covid wards and expressed anxiety at the young age of people being admitted. Other HCWs worried about exposure to Covid during unpaid hospital secondments/placements, and the risk of bringing Covid home to their own families.

#### Organisational support

HCWs acknowledged efforts to keep teams informed and promote good communication in the midst of things. However, particularly in the initial stages of the pandemic, guidelines were constantly changing, and staff received messages from multiple sources, which meant “things were blurry and confusing” (COV97, Consultant Surgeon). Uncertainty resulted in anxiety and burnout from trying to keep up with new protocols.

> *“It was really exhausting just trying to work out what we were doing with different things coming up in different places all the time. How we sort out our junior doctors, which junior doctors are staying, which are being re-deployed? You know, and so on*.

*Do we wear PPE? Do we not wear PPE?”* (COV45, Consultant, Gynecology).

Student nurses in particular, were concerned about the progress of their courses. Twitter was a platform to voice angers and concerns.

*QT @username: The silence from the NMC [Nursing and Midwifery Council] is deafening. We need clarity. Thousands of students across the country are full of anxiety waiting to hear what’s happening to them. We need guidance now @username;* □*@username if we could have clarity for all #StudentNurses ASAP it would be great. HEI’s are taking different approaches Trusts are taking different approaches #PPEroulette We want to help, but we need your *clear* guidance. We need it now. #COVID19 #RCNStudents* (COVTweet835, Student Nurse).

In some settings, work environments were more organised. HCW’s suggested that enabling direct communication channels between hospitals helped to prepare training and feel more confident, particularly communication with hospital in countries that were ahead in the pandemic curve.

> “*There was definitely a lot of planning which made everything feel very controlled. It made everything feel very calm. And it also made me feel, we felt like we knew what we were doing” (COV43, trainee GP, A&E)*.

Staff reported that strong leadership from management facilitated more compassion and awareness to mental health and wellbeing during the pandemic which they would like to be continued in future. Sentiment around the organisational support of HCW mental health was 23% negative, 70% positive and 7% neutral. Some discussion focused on the problems that HCW staff (e.g., locums and agency staff) were having accessing accommodation close to hospitals they worked in – due to hotels and other accommodation closing because of lockdown restrictions and Covid outbreaks. As well, NHS staff paid their respects to colleagues who had died from Covid-19. This led to discussions about poor staff access to PPE during the start of the pandemic, and the effects of this on staff mental health and morale. Between Feb-March 2021, some HCWs expressed relief that more people were getting their first dose of the Covid-19 vaccine, although they also warned that the UK government needed to take responsibility for failures earlier in the pandemic, as well as high levels of vaccination.

#### Social network and support

Participants highlighted family and friends as their main source of support and comfort during the peak of the pandemic, which gave them the strength to keep working in challenging conditions. The lack of social contact during the pandemic presented many challenges to some HCWs, in which they noticed a decline in their mental health as many were unable to see their family and friends for a long period of time. However, this also encouraged participants to video call their family and friends for support to maintain their wellbeing.

Sentiment with regards to support received from local social networks outside of work was 26% negative, 70% positive and 4% neutral. Some HCWs mentioned having symptoms of Long Covid (due to work-related infections) and being grateful for support of colleagues and friends who helped them to pace clinical and non-clinical hours worked. Others mentioned the positive effect of working in Covid vaccine clinics and interacting with supportive patients. While others shared tweets mentioning relief at being able to get vaccinated so that they could continue working safely.

> “*I got the Oxford AZ. My husband, also a doctor, got the Pfizer. These were the ones available to us. We didn’t care about which one, rather “thank goodness we’re vaccinated!” The #vaccine* □□*serious illness* □□*hospital admissions* □□*transmission All better than chancing COVID*□*”* (COVTwitter1277, NHS Doctor).

Some HCWs mentioned worry about working antisocial hours, and the prospect of burnout despite lower numbers of Covid patients during the summer months. Relatives of HCWs also expressed worry for the health of those who could not work from home, citing examples of hospital pharmacists whose parents had died from hospital contracted Covid, and the stress felt by HCWs that they were putting their families at risk by working on the frontline.

#### Public Support & Government support

The large majority of participants expressed keen appreciation for the positive support conveyed by the public for their efforts during the pandemic. Many staff cited the generous donations of snacks, meals, toiletries, and other personal effects as genuinely helping to sustain them during critical times. Food supplied to hospitals from restaurants and individual supporters was particularly welcomed, as it reduced the time needed for shopping/cooking and allowed more opportunity to rest after long and demanding shifts. Such outpourings of generosity were seen as buoying up the collective and individual morale, providing much needed proof that the public appreciated their hard work:

> “*The generosity of community at large outside of the hospital … the donations that we’ve had from food and all sorts of companies, to support from family and friends … and I think for me, like that’s what’s got me through this*” (COV98, Speech & Language Therapist).

The ‘Clap for Carers’ initiative was seen as a more divisive public support response. Whilst many participants appreciated the gesture, some felt it smacked of tokenism, particularly when it was juxtaposed against the flouting of social distancing rules and a seemingly blasé attitude towards the virus from some members of the public.

> *“How can people go out, stand outside on balconies, clap for frontline people and then go out and break the rules?”* (COV86, Speech & Language Therapist).

Some participants also feared that public support would gradually wane once record high waiting lists became a reality. Sentiment with regards to the perception of public support and government response was 46% negative, 48% positive and 6% neutral. While sentiment towards experience of access to PPE 45% negative, 49% positive and 6% neutral. In terms of perceptions of government support, some HCWs used twitter to express their frustration at receiving just a 1% below inflation pay rise, despite the stress and trauma experienced while working on the frontline.

> *“What a way to thank the people who’ve faced Covid straight in the eye & looked after the people suffering with it. Stayed strong to offer help to the people that are bereaved and scared. Whose job has changed so much, but we never complained. A 1% below inflation pay rise* □*”* (COVTwitter1337, Nurse Practitioner).

## DISCUSSION

### Overview of study

This study provided a though overview of HCWs experiences related to well-being during the first year of the COVID-19 pandemic. As found in the interviews and social media data, HCWs were willing and volunteered to work in unknown fields, however, redeployment generated anxiety mainly due to limited prior training and risk assessments, and the barriers of adapting to a new working environment while wearing PPE. Wellbeing support varied across hospitals and HCWs struggled to access this due to time constraints. In terms of mental health, mentions of feelings of trauma, PTSD and anxiety were prominent. In addition to the challenges of redeployment, HCWs’ mental health was particularly affected by the copious amount of bad news on media and at home and the fear of infecting their loved ones. The theme of Organizational Support exhibited dissonance in HCW experiences, with some praising the strong leadership from management whilst others explained how constantly changing guidelines caused additional anxiety.

### Redeployment, clinical work, and sense of duty

Similarly, to other studies in the UK, we found that redeployment generated anxiety for staff [4, 35] Our results did not capture the impact of redeployment to specific areas of the healthcare system, which is seemingly a major contributor to poor mental health in staff. However, other studies consistently found redeployment specifically to ICU units caused the most depressive symptoms and subsequently had the greatest impact on the mental health of HCWs in the UK [36, 37]. This study showed evidence that during the COVID-19 pandemic this was likely due to feeling ill equipped, receiving limited training, and limited room for informal interactions within and outside clinical settings. Recent evidence supports this by indicating that the quantity and quality of training received prior to redeployment has a strong impact on HCW’s mental health [37]. This study showed evidence that during the COVID-19 pandemic this was likely due to feeling ill equipped, receiving limited training, and limited room for informal interactions within and outside clinical settings. Recent evidence supports this by indicating that the quantity and quality of training received prior to redeployment has a strong impact on HCW’s mental health [37]. Other studies emphasise that feeling ill equipped is most strongly associated with shortages of PPE [38, 39].

### Mental health effects & Well-being support, coping strategies

As has been reported elsewhere, HCWs reported a wide range of mental health effects of delivering care during the pandemic [40-43]. These included PTSD, trauma, and exhaustion. It has been well documented that these feelings are more pronounced in ICU workers, due to high workloads, daily exposure to death and irregular working hours [44]. Even in non-emergency situations, ICU workers suffer from the highest levels of anxiety compared to other units [45]. These results, together with the results from our study, suggest that this line of research is relevant beyond the context of the pandemic, and there is sufficient information to move on to planning strategies to address this issue.

In terms of coping with these damaging mental health effects, HCWs perceptions of the support available to them were mixed. Similar to Schecter et al [46] some found sessions with clinical psychologists helpful, while others indicated that the wellbeing support offered at their hospitals did not align to their needs or work patterns. HCWs often appreciated and relied more on informal support from family friends and colleagues [25, 47]. Muller et al [24] mirrored this idea by demonstrating that addressing organizational factors and support would improve mental health and well-being more than psychological help.

Also, some HCWs stated that they used their own methods to cope, such as mindfulness apps, with growing evidence that mindfulness-based apps improve wellbeing [48]. Meanwhile, in New York, physical activity appeared to be the popular coping mechanism [46].

### Organisational support

A systematic review concluded that mental health problems in HCWs during COVID-19 correlated with the organizational failures of the healthcare system [24]. Our results support this finding as organizational support emerged as a main theme in discussions with HCWs. Mostly, HCWs deemed leadership from management as strong and that it facilitated compassion for mental health and well-being. However, this was overshadowed by the constantly changing guidelines, which led to uncertainty. This has been confirmed by other studies [4], where uncertain and changing guidelines, as well as the backlog of patient care, caused superfluous cognitive burdens. A systematic review conducted by our team identified key principles for successful redeployment based on a compilation of strategies applied around the world [49]. These included, for example, providing repeated shorter practical training sessions, rather than receiving all information in one session which is less flexible to adapt to emerging content and staff availability.

Additionally, some studies found that HCWs felt supported by hospital administration but not by their supervisors [35]. Our study provided insight into excessive responsibilities of managers that prevented them from carrying out their role as they would have liked, particularly in the case of nurses who were significantly understaffed. Improvements in the organization of care and a clearer sense of guidelines also reassured staff.

### Social network & Public support & Government support

In terms of support from their social networks, the public and the government, HCWs were grateful and appreciative for their families and friends. The lack of socialization caused further damage to their well-being, as was seen for other populations during these times. It appears that social support is an integral element of maintaining HCWs mental health as previous studies have found that a lack of social support affects sleep, anxiety, and stress [50]. In our previous related study [4] we also found that HCW placed significant emphasis on their responsibilities towards loved ones who consistently supported them, and the invaluable support of the community. This was mirrored by Sun et al [51] who identified that team support was a major protective factor against poor mental health during the pandemic.

In regard to public support, HCWs were appreciative of the donations, meals prepared and support from the public. Such outpourings of generosity were seen as buoying up the collective and individual morale and positive feelings about the public appreciating their hard work. Although, it was sometimes deemed tokenistic, when HCWs compared their hardships to the large number of members of the public regularly breaking COVID-19 guidelines. These mixed perceptions on public support were supported by the sentiment analysis, which found a roughly equal amount of positive and negative tweets concerning the topic. A topic that did not come up in this study was HCWs of Asian origin feeling discriminated against by the public [52]. This study perhaps did not pick up on this due to a lack of diversity within the sample, as the majority of participants were of White British background.

### Deep-rooted attitude: fears of infecting families

HCWs fear and anxiety of passing on COVID-19 to their families is what is known as a deep-rooted attitude in qualitative research [53]. as it permeated all aspects of their descriptions around wellbeing. The damaging mental health effects that HCWs stated they felt were centered around the prospect of infecting others and being the cause of their suffering. When discussing support from their social networks, this idea persisted as participants mentioned that they could not fully reap the benefits of their loved ones’ support due to this anxiety of transmitting the virus. PPE shortages generated concern in other studies too due to the possibility of getting COVID-19 and passing it on to family members [8, 39]. This finding is not new, as it has been seen in past pandemics as well [54], emphasising the need to include it as a priority in emergency response guidelines.

### Limitations

The study had several limitations. The HCWs included in the interview study worked in London, limiting the representation of experiences in other areas of the country, and the generalizability of our findings. However, HCWs perceptions were captured through interviews and social media too which helps to mitigate this limitation. Even though we collected data over 12 months, the last interviews were carried out in March 2021, limiting our exploration of experiences in later stages of the pandemic. In this regard, one study concluded that anxiety levels in HCWs have increased since the pandemic was declared [55].

Although purposive sampling was used to obtain a varied sample in terms of gender, role and ethnicity, most interview participants were of White British backgrounds, and it is not possible to determine diversity for the social media sample. HCWs of different ethnic backgrounds are likely to have had different experiences and perceptions to share and further research looking specifically at their perspectives should be conducted.

### Implications & recommendations for future research

The psychological reactions of the population during an infectious disease pandemic play a crucial role in determining not only the progression of the pandemic but also the wellbeing during and after peaks of infection. This is even more important to consider for HCWs, as they are at the forefront of the battle. The World Health Organisation is officially acknowledging this risk to HCWs mental health, stating that additional long-term support must be provided, to prevent symptoms of PTSD and depression [56]. Notwithstanding this, limited resources are allocated to HCWs broader wellbeing, as efforts focus mainly on mitigating potential clinical mental health problems.

The results of this study emphasise the need for considering HCWs personal perspective to understand their context and barriers and facilitators to wellbeing. Although previous studies have recommended tailored psychological interventions to manage symptoms and consolidate appropriate coping strategies, the results of this study imply that more systemic action should be taken [20, 57]. For instance, encouragement of HCW-led wellbeing initiatives such as You ok, Doc? [58] Doctors Care [59], and <u>EveryDoctor [60].</u> This could facilitate more open conversations between peers, sharing of experiences and coping strategies outside of clinical settings, as well as empower HCWs to inform the shape of health service provision. Another way in which this could be achieved is by creating a more comfortable environment for HCWs, having adequate and appropriate equipment, most importantly, PPE, increased staff testing, to ease the anxiety of transmitting the virus to family members, could also address mental health and wellbeing concerns. As described above, these aspects are for the most part overlooked as necessary forms of staff wellbeing support that hospital administration, with support from the government, should consider.

Future research should investigate personal factors which could potentially put some HCWs more at risk of damaging mental health and well-being effects. This is because Yao et al [36] concluded that women, individuals who were married and those who had engaged in less than 7 years of clinical work, were more negatively affected by the COVID-19 pandemic. Wozniak et al [44] supported these conclusions but also demonstrated that PHQ-9 and GAD-7, assessments of symptoms of depression and different types of anxiety, were worse for those working in ICU versus those in non-ICU settings. In addition, while this study tried to include a broad sample of frontline HCWs, other hospital staff such as professional services, security, and cleaning staff who struggled with the immense workload and stress were not possible to reach often because they are not direct NHS employees. Obtaining their perspective is essential to fully understand the dynamics and perceptions about wellbeing in healthcare services.

### Conclusion

Our results provide a clear depiction of HCWs personal perspective of wellbeing needs, experiences and strategies used to maintain wellbeing. Our findings make evident the need for a systemic approach, broader than the current clinical focus on mental health problems. These findings should be used to inform future pandemic response guidelines and wellbeing funding allocation decisions. Additionally, the innovative method applied in this study has proved to yield useful results for applied research in an efficient manner. This method is recommended for future emergency response research looking at big qualitative data.

## Data Availability

Data included in this study is not available outside the Rapid Research Evaluation and Appraisal Lab (RREAL) due to ethical guidelines restrictions.

## Acknowledgements

NV, SM, and CV led the conception, secondary data gathering, analysis, drafting and final revision of the paper.

AB, LM, PCG, CB, ECF, SV, AS, SMS and IU were involved in data collection, analysis, drafting and revision of the paper.

AK, SI were involved in consolidating results, drafting, and revision of the paper.

We acknowledge the contributions of Dr Ginger Johnson, who provided general supervision of the larger research project.

The study did not receive external funding. Research costs were covered with existing funding provided by the Rapid Research Evaluation and Appraisal Lab (RREAL), University College London. All researchers acted independently from the funding source and can take responsibility for the integrity of the data and the accuracy of the data analysis.

## Abbreviations

HCW: Health Care Worker
PPE: Personal Protective Equipment
SPICE: Recovery Model of Social recovery, Prosperity, Individual recovery and Clinical recovery experience
RREAL: Rapid Research Evaluation and Appraisal Lab
LISTEN: Collaborative and Digital Analysis of Big Qualitative Data in Time Sensitive Contexts
HRA: Health Research Authority

## Appendices

### Appendix 1. Boolean search terms

#### Mental health effects

((bio:”healthcare professional” **OR** bio:”healthcare worker” **OR** bio:”doctor” **OR** bio:”GP” **OR** bio:”Pharmaci*” **OR** bio:”Radiograph*” **OR** bio:”Therap*” **OR** bio:”Neurolo*” **OR** bio:”Pyschol*” **OR** bio:”clinic*” **OR** bio:”nurse” **OR** bio:”physio*” **OR** bio:”midwi*” **OR** bio:”obstet*” **OR** bio:”geriatr*”) **AND** (“coronavirus” **OR** “#coronavirus” **OR** “corona” **OR** “COVID-19” **OR** “COVID 19” **OR** “COVID19” **OR** “#COVID19” **OR** “COVID_19” **OR** “COVID”) **AND** (“redeploy*” **OR** “stress*” **OR** “overwork*” **OR** “face-to-face” **OR** “face to face” **OR** “anxiet*” **OR** “anxi*” **OR** “scared” **OR** “afraid” **OR** “tired” **OR** “trauma*” **OR** “burn* out” **OR** “burnout” **OR** “not able” **OR** “insomni*” **OR** “sad*” **OR** “bad news” **OR** “mental health” **OR** “fright*” **OR** “worr*” **OR** “scared” **OR** “afraid” **OR** “tired” **OR** “alcho*” **OR** “sleep*” **OR** “rest*” **OR** “drink*” **OR** “alcohol*” **OR** “drug*” **OR** “smok*” **OR** “dying*” **OR** “death*” **OR** “personal” **OR** “pending” **OR** “expect*” **OR** “detach*” **OR** “autopilot” **OR** “auto pilot” **OR** “cope*” **OR** “coping” **OR** “responsi*” **OR** “access*” **OR** “support*” **OR** “friend*” **OR** “adequa*” **OR** “vacci*” **OR** “lockdo*” **OR** “first wave*” **OR** “risk*” **OR** “unfamilia*” **OR** “second wave*” **OR** “firstwave*” **OR** “secondwave*”))

#### Organisational

((bio:”healthcare professional” **OR** bio:”healthcare worker” **OR** bio:”doctor” **OR** bio:”GP” **OR** bio:”Pharmaci*” **OR** bio:”Radiograph*” **OR** bio:”Therap*” **OR** bio:”Neurolo*” **OR** bio:”Pyschol*” **OR** bio:”clinic*” **OR** bio:”nurse” **OR** bio:”physio*” **OR** bio:”midwi*” **OR** bio:”obstet*” **OR** bio:”geriatr*”) **AND** (“coronavirus” **OR** “#coronavirus” **OR** “corona” **OR** “COVID-19” **OR** “COVID 19” **OR** “COVID19” **OR** “#COVID19” **OR** “COVID_19” **OR** “COVID”) **AND** (“at work” **OR** “hospital” **OR** “clinic*” **OR** “ward*” **OR** “ICU*” **OR** “unit*” **OR** “NHS trust*”) **AND** (“co-worker*” **OR** “co worker*” **OR** “communi*” **OR** “comms” **OR** “colleague*” **OR** “team*” **OR** “group*” **OR** “wellbeing” **OR** “leader*” **OR** “staff*” **OR** “dynam*” **OR** “manag*” **OR** “redploy*” **OR** “deploy*” **OR** “access*” **OR** “unfair*” **OR** “mental health*” **OR** “mentalhealth*” **OR** “ppe*” **OR** “clear*” **OR** “clarity” **OR** “anxi*” **OR** “worr*” **OR** “guide*” **OR** “polici*” **OR** “policy*” **OR** “compulso*” **OR** “voluntar*” **OR** “duties*” **OR** “duty” **OR** “profression*” **OR** “organisat*” **OR** “manag*” **OR** “upskill*” **OR** “skill*” **OR** “network*” **OR** “abilit*” **OR** “develop*” **OR** “remote*” **OR** “isolat*”))

#### Social network/ support network

((bio:”healthcare professional” **OR** bio:”healthcare worker” **OR** bio:”doctor” **OR** bio:”GP” **OR** bio:”Pharmaci*” **OR** bio:”Radiograph*” **OR** bio:”Therap*” **OR** bio:”Neurolo*” **OR** bio:”Pyschol*” **OR** bio:”clinic*” **OR** bio:”nurse” **OR** bio:”physio*” **OR** bio:”midwi*” **OR** bio:”obstet*” **OR** bio:”geriatr*”) **AND** (“coronavirus” **OR** “#coronavirus” **OR** “corona” **OR** “COVID-19” **OR** “COVID 19” **OR** “COVID19” **OR** “#COVID19” **OR** “COVID_19” **OR** “COVID”) **AND** (“family*” **OR** “friend*” **OR** “support*” **OR** “relationsh*” **OR** “interact*” **OR** “help*” **OR** “group*” **OR** “partner*” **OR** “daughter*” **OR** “wife*” **OR** “husband*” **OR** “boyfriend*” **OR** “bff” **OR** “girlfriend*” **OR** “son*” **OR** “mum*” **OR** “mother*” **OR** “father*” **OR** “mate*” **OR** “dad*”))

#### Wider context

((bio:”healthcare professional” **OR** bio:”healthcare worker” **OR** bio:”doctor” **OR** bio:”GP” **OR** bio:”Pharmaci*” **OR** bio:”Radiograph*” **OR** bio:”Therap*” **OR** bio:”Neurolo*” **OR** bio:”Pyschol*” **OR** bio:”clinic*” **OR** bio:”nurse” **OR** bio:”physio*” **OR** bio:”midwi*” **OR** bio:”obstet*” **OR** bio:”geriatr*”) **AND** (“coronavirus” **OR** “#coronavirus” **OR** “corona” **OR** “COVID-19” **OR** “COVID 19” **OR** “COVID19” **OR** “#COVID19” **OR** “COVID_19” **OR** “COVID”) **AND** (“public*” **OR** “support*” **OR** “response*” **OR** “government*” **OR** “measures” **OR** “disillus*” **OR** “accommodat*” **OR** “living arrangement*” **OR** “government respons*” **OR** “government measure*” **OR** “impact*” **OR** “public support*” **OR** “clapforcarers*” **OR** “clap for carers*” **OR** “clap-for-carer*” **OR** “rainbow*” **OR** “nhs rainbow*” **OR** “childcarei*”))

#### PPE

((bio:”healthcare professional” **OR** bio:”healthcare worker” **OR** bio:”doctor” **OR** bio:”GP” **OR** bio:”Pharmaci*” **OR** bio:”Radiograph*” **OR** bio:”Therap*” **OR** bio:”Neurolo*” **OR** bio:”Pyschol*” **OR** bio:”clinic*” **OR** bio:”nurse” **OR** bio:”physio*” **OR** bio:”midwi*” **OR** bio:”obstet*” **OR** bio:”geriatr*”) **AND** (“coronavirus” **OR** “#coronavirus” **OR** “corona” **OR** “COVID-19” **OR** “COVID 19” **OR** “COVID19” **OR** “#COVID19” **OR** “COVID_19” **OR** “COVID”) **AND** (“PPE*” **OR** “ppe*”) **AND** (“physical” **OR** “breath*” **OR** “hot*” **OR** “anxiet*” **OR** “scared” **OR** “afraid” **OR** “tired” **OR** “abilit” **OR** “access*” **OR** “ramadan” **OR** “eid” **OR** “eid al-adha” **OR** “dhydrat*” **OR** “hydrat*” **OR** “slow*” **OR** “headach*” **OR** “mood*” **OR** “emotion*” **OR** “worr*” **OR** “perform*” **OR** “usual*” **OR** “tired” **OR** “wellbeing” **OR** “worr*” **OR** “sweat*” **OR** “group*” **OR** “professi*” **OR** “comms*” **OR** “communic*” **OR** “speak*” **OR** “hear*” **OR** “clear*” **OR** “clinic*”))

### Appendix 2. Sentiment analysis

#### Sentiment analysis criteria for mental health and wellbeing of HCWs

HCW experience of mental health and wellbeing during Covid-19 pandemic

#### Definition & Context

We aim to gather accounts of the experiences of healthcare workers (HCWs) in the challenges and constraints they might have to their mental health and wellbeing during the COVID-19 pandemic.

#### Sentiment analysis of mental health and wellbeing

##### Positive (P)

- Post communicating overall trust and satisfaction with experience of or guidelines and support for mental health and wellbeing in the context of the COVID-19 pandemic
- Posts are affirming of mental health and wellbeing delivery and experiences of staff

##### Negative (N)

- Post contains negative attitude/arguments against Covid-19 treatment / guidelines / support / of HCW mental health and wellbeing
- Post discourages the following of recommended treatment / guidelines / support related to mental health and wellbeing
- Post shares bad HCW experiences of working on Covid frontline and the effect of this on mental health and wellbeing.

##### Neutral (NT)

- Post contains no elements of uncertainty, positive or negative content.
- Post contains general statement(s) or link(s) to item(s) (e.g., news articles/papers) with no expression of sentiment.
- Post includes factual statements/recommendations about COVID-19 and mental health and wellbeing, but no other sentiment.

### Appendix 4. Social media analysis: Inclusion and exclusion criteria

| Inclusion Criteria Definition |
| --- |
| Content refers specifically to the experience of HCWs within the overall topic of mental health and wellbeing as covered in the interview analysis. I.e., People actually talking about wellbeing topics, rather than things we infer might affect wellbeing (without them mentioning it). |
| Example of tweet to include: "Government is not supporting healthcare workers." We would include this because it mentions explicitly about HCWs, something that is affecting HCWs (and therefore their wellbeing) directly. |
| Exclusion Criteria Definition |
| Content does NOT refer specifically to the experience of HCWs within the overall topic of mental health and wellbeing. |
| Example to exclude: "I am against government budget cuts". This is something that ultimately probably affects healthcare workers, but there's no explicit mention to them or their wellbeing. Even if it's a HCW writing it, we can't know if they're against government cuts because of something work/wellbeing-related |

